# METFORMIN USE IS ASSOCIATED WITH REDUCED MORTALITY IN A DIVERSE POPULATION WITH COVID-19 AND DIABETES

**DOI:** 10.1101/2020.07.29.20164020

**Authors:** Andrew B. Crouse, Tiffany Grimes, Peng Li, Matthew Might, Fernando Ovalle, Anath Shalev

## Abstract

**BACKGROUND:** Coronavirus disease-2019 (COVID-19) is a growing pandemic with an increasing death toll that has been linked to various comorbidities as well as racial disparity. However, the specific characteristics of these at-risk populations are still not known and approaches to lower mortality are lacking.

**METHODS:** We conducted a retrospective electronic health record data analysis of 25,326 subjects tested for COVID-19 between 2/25/20 and 6/22/20 at the University of Alabama at Birmingham Hospital, a tertiary health care center in the racially diverse Southern U.S. The primary outcome was mortality in COVID-19-positive subjects and the association with subject characteristics and comorbidities was analyzed using simple and multiple linear logistic regression.

**RESULTS:** The odds ratio of contracting COVID-19 was disproportionately high in Blacks/African-Americans (OR 2.6; 95%CI 2.19-3.10; p<0.0001) and in subjects with obesity (OR 1.93; 95%CI 1.64-2.28; p<0.0001), hypertension (OR 2.46; 95%CI 2.07-2.93; p<0.0001), and diabetes (OR 2.11; 95%CI 1.78-2.48; p<0.0001). Diabetes was also associated with a dramatic increase in mortality (OR 3.62; 95%CI 2.11-6.2; p<0.0001) and emerged as an independent risk factor in this diverse population even after correcting for age, race, sex, obesity and hypertension. Interestingly, we found that metformin treatment was independently associated with a significant reduction in mortality in subjects with diabetes and COVID-19 (OR 0.33; 95%CI 0.13-0.84; p=0.0210).

**CONCLUSION:** Thus, these results suggest that while diabetes is an independent risk factor for COVID-19- related mortality, this risk is dramatically reduced in subjects taking metformin, raising the possibility that metformin may provide a protective approach in this high risk population.

## INTRODUCTION

Coronavirus disease-2019 (COVID-19) caused by the severe acute respiratory syndrome-coronavirus-2 (SARS-CoV-2) is a growing global pandemic that has devastated Asia, Europe and now the United States. Its increasing death toll has been linked to higher age and a number of comorbidities including hypertension, obesity and diabetes (1, 2), but approaches to counteract this trend are still lacking. Being a new disease, the specific patient characteristics of these at risk populations are also only starting to emerge with studies reported from China (3–5), Europe (6, 7) and more recently New York (1, 2).

However, currently still very little is known about patient characteristics in the U.S., particularly in more diverse communities with a large proportion of Blacks/African-Americans such as in the South. This information is especially relevant as African-Americans have been disproportionally affected by this pandemic across the nation (8–10) and the prevalence of comorbidities including diabetes is very high in these communities (11). We therefore conducted a retrospective observational study of subjects diagnosed with COVID-19 at the University of Alabama at Birmingham (UAB) Hospital, a tertiary health care center in the South, aimed at identifying the patient characteristics and factors affecting mortality especially in the context of diabetes in this diverse cohort.

## METHODS

### Study design and participants

We conducted a retrospective analysis of de-identified electronic health record data from subjects consecutively tested for COVID-19 between February 25, 2020 and June 22, 2020 at UAB (Institutional Review Board protocol E160105006). Subjects were categorized as confirmed COVID-19 positive or negative based on RT-PCR results from SARS-CoV-2 viral nucleic acid testing in respiratory specimens. The primary outcome was mortality and the effects of patient characteristics and comorbidities were analyzed.

### Statistical Analysis

Patient characteristics and comorbidities were summarized as mean and standard deviation (SD) for continuous variables; and frequency and proportion for categorical variables. In analysis, age was categorized into three groups: <50, 50-70, and >70 years old. The association with COVID-19 diagnosis was explored utilizing a simple linear logistic regression for each of the potential risk factors and the raw odds ratio (OR) and the 95% confidence interval (95%CI) were calculated for the strength of association. The associations with COVID- 19 mortality for the potential risk factors were explored with both simple linear logistic regression for raw ORs and multiple linear logistic regression for adjusted ORs. Potential interactions were evaluated and removed from the multiple logistic regression model if not significant. All the tests were two-sided under the significance level of 0.05. The analyses were conducted using SAS 9.4 (Cary, NC).

## RESULTS

### Subject characteristics and COVID-19 diagnosis

The characteristics of the 24,722 subjects who tested negative for COVID-19 and 604 subjects who had a confirmed positive COVID-19 test are listed in **Table 1**. To explore the association between COVID-19 diagnosis and potential risk factors, a simple logistic regression was used. Notably, despite only representing 26% of the population in Alabama, the number of African-Americans who tested positive for COVID-19 was disproportionally high as African-Americans represented 52% of those who tested positive while accounting for only 30% of those who tested negative. This resulted in a highly significant odds ratio (OR 2.6, 95%CI 2.19-3.10; p<0.0001) **(Table 1)**. In contrast, only 36% of COVID-19 positive subjects were white, whereas Whites made up 56% of those who tested negative, further underlining the racial disparity. Interestingly, 70% of all subjects diagnosed with COVID-19 had pre-existing hypertension, 61% had obesity and 40% diabetes and the risk of being diagnosed with COVID-19 while suffering from any one of these comorbidities was significantly elevated (p<0.0001) **(Table 1)**. These results are very much in line with global observations and suggested that our cohort provided a representative sample.

**Table 1:** Subject characteristics and COVID-19 diagnosis.

| Subject characteristics | COVID-19 |  | Comparison | OR (95%CI) | P-value |
| --- | --- | --- | --- | --- | --- |
|  | Negative<br>(n=24,722) | Positive<br>(n=604) |  |  |  |
| Age Group |  |  |  |  |  |
| <50 years | 10626 (43.0%) | 239 (39.6%) |  |  |  |
| 50-70 years | 9862 (39.9%) | 245 (40.6%) | 50-70 vs <50 | 1.10 (0.92, 1.32) | 0.2798 |
| >70 years | 4234 (17.1%) | 120 (19.9%) | >70 vs 50-70 | 1.14 (0.91, 1.42) | 0.2432 |
| Race |  |  |  |  |  |
| African-American (AA) | 7498 (30.3%) | 311 (51.5%) | AA vs White | 2.61 (2.19, 3.10) | <0.0001 |
| White | 13821 (55.9%) | 220 (36.4%) |  |  |  |
| Other | 3403 (13.8%) | 73 (12.1%) |  |  |  |
| Sex |  |  |  |  |  |
| Male | 10671 (43.2%) | 272 (45.0%) | M vs F | 1.06 (0.90, 1.25) | 0.4629 |
| Female | 13841 (56.0%) | 332 (55.0%) |  |  |  |
| Unidentified | 210 (0.8%) |  |  |  |  |
| Obesity |  |  |  |  |  |
| Yes | 11167 (45.2%) | 371 (61.4%) | Y vs N | 1.93 (1.64, 2.28) | <0.0001 |
| No | 13555 (54.8%) | 233 (38.6%) |  |  |  |
| Hypertension |  |  |  |  |  |
| Yes | 11891 (48.1%) | 420 (69.5%) | Y vs N | 2.46 (2.07, 2.93) | <0.0001 |
| No | 12831 (51.9%) | 184 (30.5%) |  |  |  |
| Diabetes |  |  |  |  |  |
| Yes | 5865 (23.7%) | 239 (39.6%) | Y vs N | 2.11 (1.78, 2.48) | <0.0001 |
| No | 18857 (76.3%) | 365 (60.4%) |  |  |  |

### Characteristics and mortality of COVID-19 positive subjects

Overall mortality in COVID-19 positive individuals was 11%, but varied a lot depending on a number of subject characteristics. 93% of deaths occurred in subjects over the age of 50 and male sex as well as hypertension were associated with a significantly elevated risk of death as assessed by bivariate logistic regression analysis **(Table 2)**. In addition, diabetes was associated with a dramatic increase in mortality (OR 3.62; 95%CI 2.11-6.2; p<0.0001). In fact, 67% of deaths occurred in subjects with diabetes.

**Table 2:**
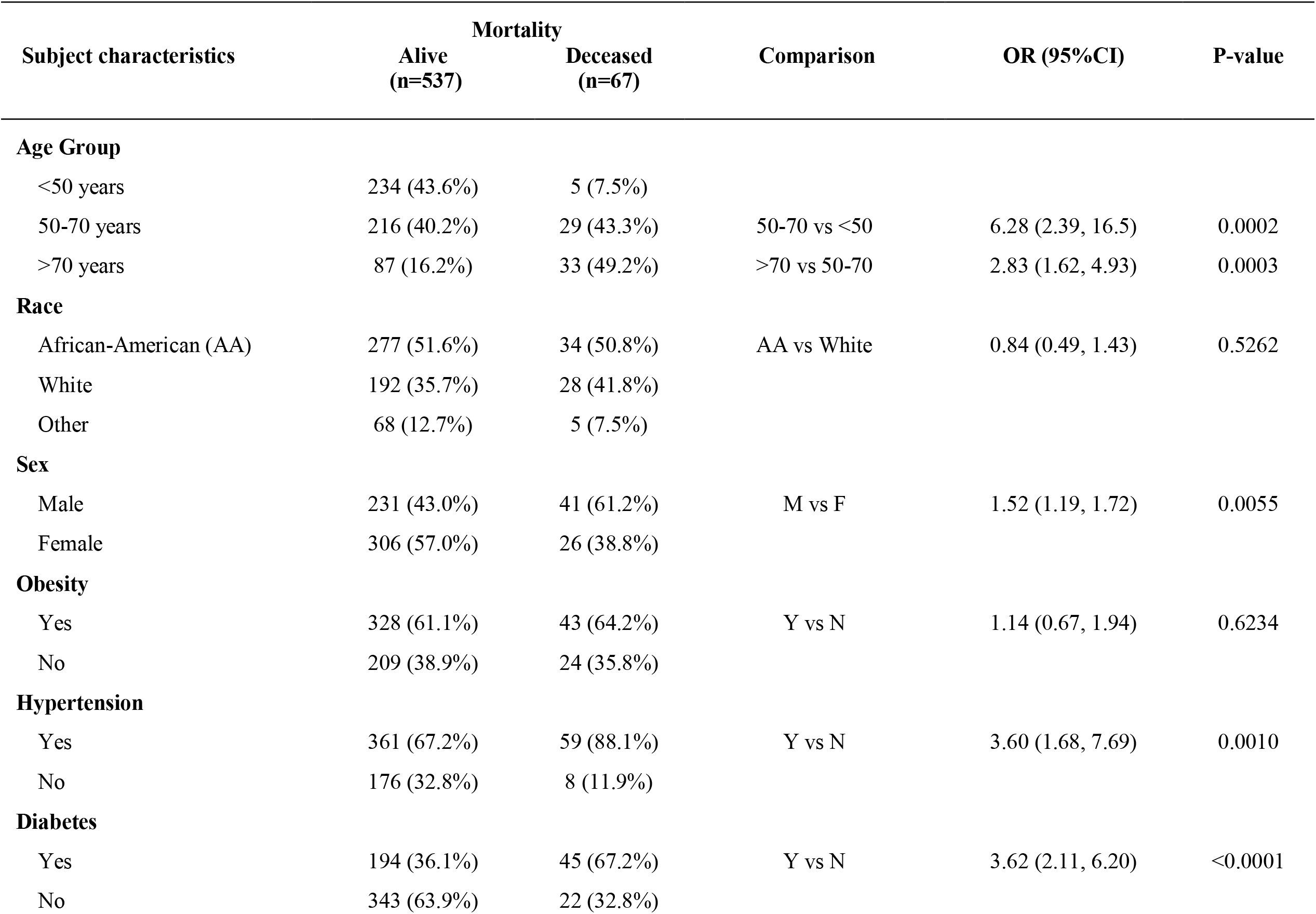
Characteristics and mortality of COVID-19 positive subjects.

We also conducted multiple logistic regression analysis with age, race, sex, obese status, hypertension status, and diabetes status as covariates and the adjusted odds ratios and 95%CIs are illustrated in **Figure 1**. Specifically, after controlling for these other covariates, age, sex, and diabetes emerged as the major factors significantly associated with COVID-19 related mortality, suggesting that they are independent risk factors.

**Figure 1.**
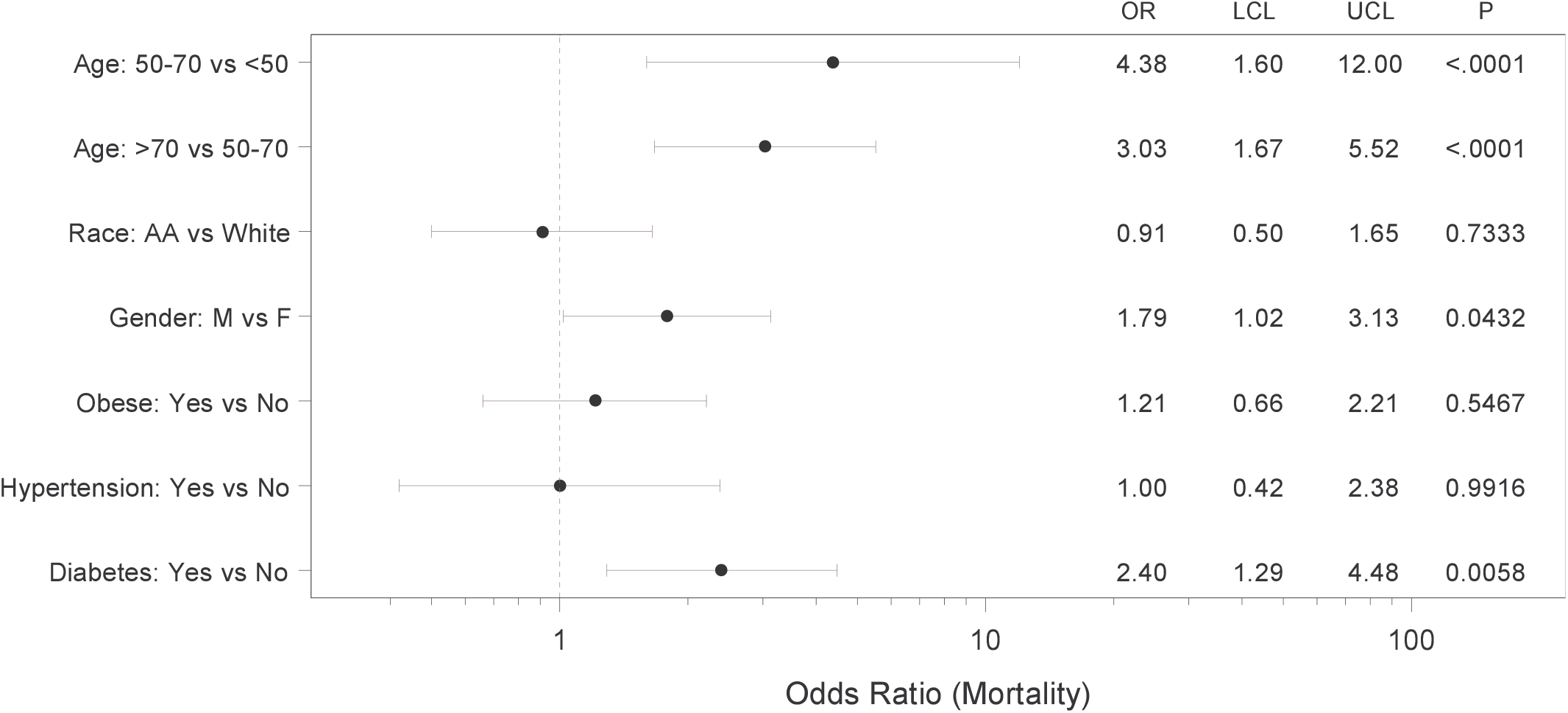
Forest plot showing adjusted mortality risk in subjects with COVID-19. Multiple logistic regression analysis with age, race, sex, obese status, hypertension status, and diabetes status as covariates was performed. The regression yielded a significant model (p<0.0001) with AUC of 0.79 (95%CI 0.74-0.85) and the adjusted odds ratios (OR), 95% confidence intervals (LCL - UCL) and corresponding P-values are shown.

### Characteristics and mortality of COVID-19 positive subjects with diabetes

Based on the identification of diabetes as an independent risk factor for mortality in COVID-19 positive subjects, we explored potential additional risk factors within this diabetic subgroup. Notably, higher age and male sex continued to be associated with increased mortality in the context of diabetes, while no significant difference between type 1 (T1D) and type 2 diabetes (T2D) was observed **(Table 3)**. Next, we investigated the effects of diabetes treatment on adverse COVID-19 outcome. We focused on insulin and metformin as the two most common medications prescribed for T2D. To avoid confounding effects from insulin being initiated for stress hyperglycemia and from metformin being discontinued in hospitalized patients, only medications used prior to the diagnosis of COVID-19 were considered. Interestingly, while prior insulin use did not seem to affect mortality risk, metformin use significantly reduced the odds of dying (OR 0.38; 95%CI 0.17-0.87; p=0.0221). In fact, with 11% the mortality of metformin users was comparable to that of the general COVID-19-positive population and dramatically lower than the 23% mortality observed in subjects with diabetes and not on metformin. Of note, this beneficial effect of metformin use on adverse outcome remained even when subjects with chronic kidney disease or chronic heart failure, classical contraindications for metformin, were excluded from the analysis (OR 0.17; 95%CI 0.04-0.79; p=0.0231). This makes any potential confounding effects from skewing metformin users towards healthier subjects without these additional comorbidities, very unlikely.

**Table 3:**
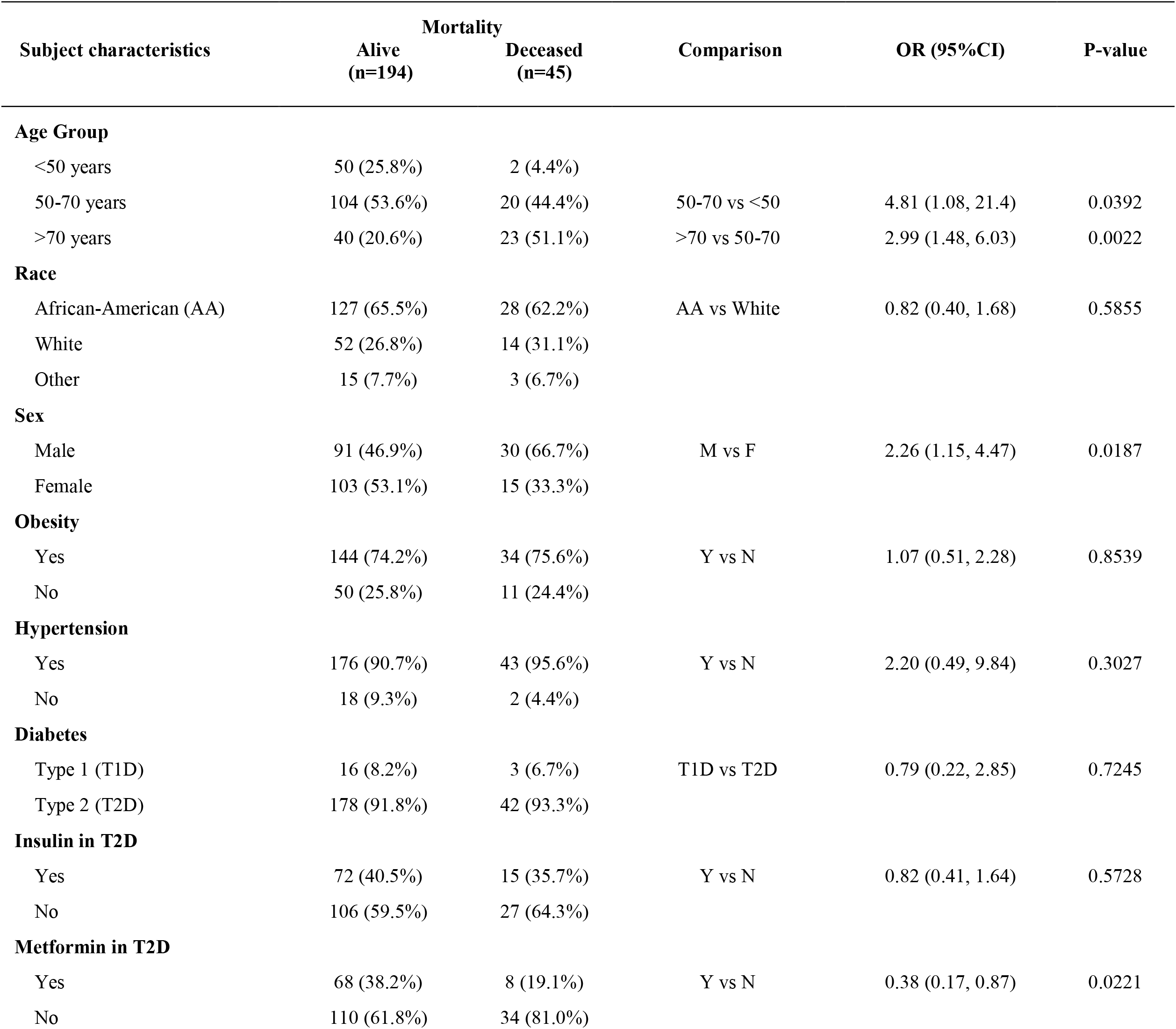
Characteristics and mortality of COVID-19 positive subjects with diabetes.

Moreover, we again performed multiple logistic regression analysis with metformin use, insulin use, age, race, sex, obese status, and hypertension status as covariates and the adjusted odds ratios and 95%CIs are shown in **Figure 2**. Specifically, after controlling for other covariates, age, sex and metformin use emerged as independent factors affecting COVID-19 related mortality. Interestingly, even after controlling for all these other covariates, the likelihood of death for subjects taking metformin for their T2D was significantly less than for those who did not take metformin (OR 0.33; 95%CI 0.13-0.84; p=0.0210).

**Figure 2.**
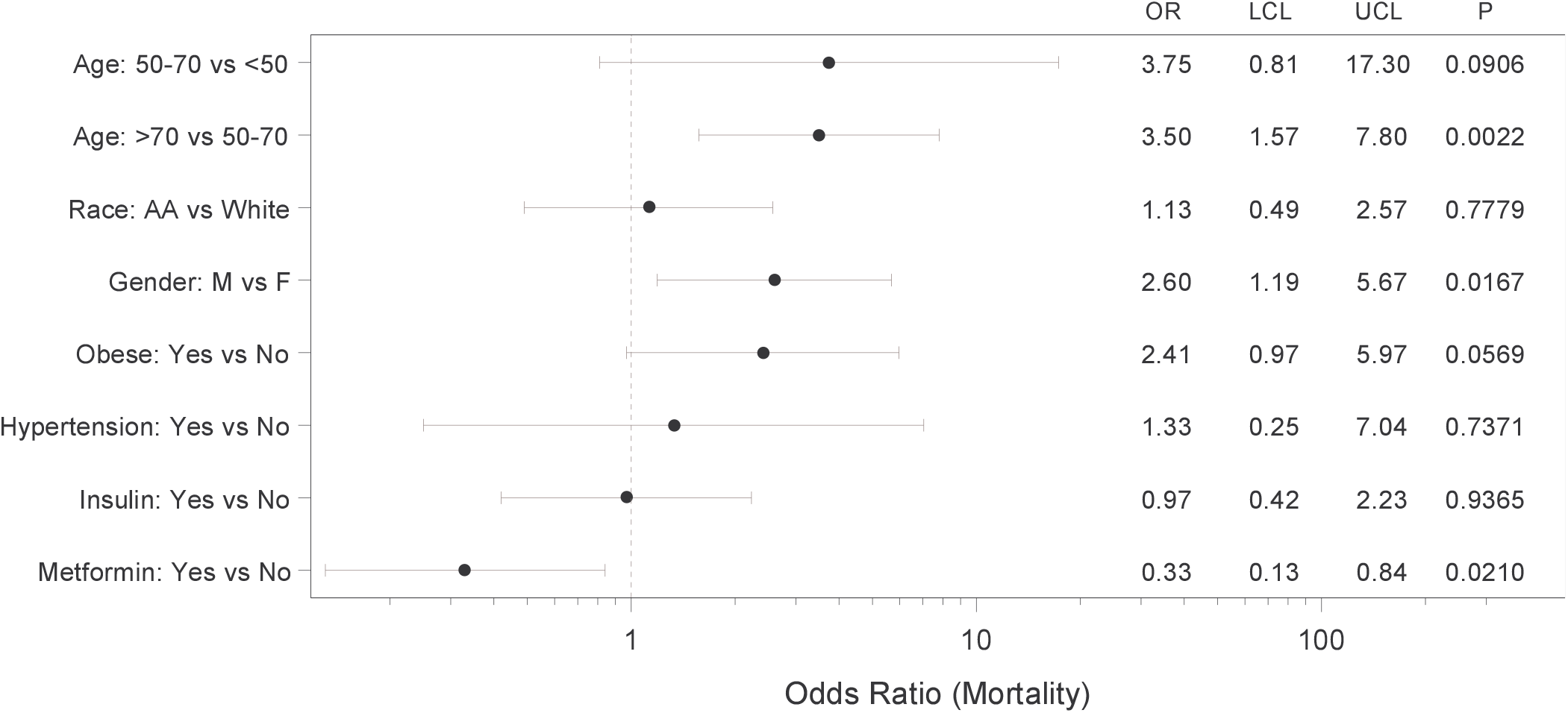
Forest plot showing adjusted mortality risk in subjects with COVID-19 and T2D. Multiple logistic regression analysis with metformin use, insulin use, age, race, sex, obese status, and hypertension status as covariates was performed and yielded a significant model (p=0.0001) with AUC of 0.77 (0.69, 0.85). The adjusted odds ratios (OR), 95% confidence intervals (LCL - UCL) and corresponding P-values are shown.

Since metformin is known and used for its weight neutral or even weight lowering properties, while improving glycemic control in T2D (12), we wondered whether these effects might explain its association with reduced risk of COVID-19 related mortality. However, neither body mass index (BMI) nor hemoglobin A1C (HbA1C) were lower in metformin users who survived as compared to those who died **(Table 4)**. While surprising, this is consistent with the notion that long-term glycemic control does not affect COVID-19 outcome, as recently reported (6). Furthermore, it suggests that other factors may play a more important role in terms of outcome in the context of COVID-19, T2D and metformin.

**Table 4:** BMI and HbA1C of subjects with COVID-19 and T2D treated with/without metformin.

|  |  | Alive |  | Deceased |  | P-value |
| --- | --- | --- | --- | --- | --- | --- |
|  |  | Mean | SD | Mean | SD | *t-test* |
| **BMI (kg/m<sup>2</sup>)** | Metformin use - Yes | 35.2 | 9.4 | 30.9 | 6.9 | 0.2406 |
|  | Metformin use - No | 33.6 | 8.7 | 32.4 | 9.8 | 0.6039 |
| **HbA1C (%)** | Metformin use - Yes | 8.0 | 2.6 | 7.3 | 1.3 | 0.4627 |
|  | Metformin use - No | 7.0 | 1.8 | 6.6 | 2.1 | 0.4428 |

## DISCUSSION

In summary, the findings of this study demonstrate that diabetes is an independent risk factor associated with increased mortality in individuals with COVID-19, whereas metformin treatment is associated with dramatically reduced mortality in subjects with T2D even after correcting for multiple covariates.

Limitations of the study include the size that did not allow for any separate analyses of additional subgroups such as T1D or subjects on other anti-diabetic drugs besides metformin. On the other hand, the diverse community comprising a large proportion of African-American men and women represents a unique feature of our study.

Most strikingly, we found that metformin use prior to the diagnosis of COVID-19 was associated with a ~3-fold decrease in mortality and significantly lower unadjusted and adjusted odds ratios in subjects with diabetes. Of note, this effect remained even after correcting for age, sex, race, obesity and hypertension or chronic kidney disease and heart failure. Interestingly and in alignment with this finding, an early report from Wuhan, China also suggested that metformin was associated with decreased mortality in hospitalized COVID-19 patients with diabetes in (13). Metformin was also found to be associated with reduced risk of early death in the French CORONADO study (6) and most recently, it was suggested to be associated with decreased mortality in women with COVID-19 based on a UnitedHealth data analysis (14). The fact that such similar results were obtained in different populations from around the world suggests that the observed reduction in mortality risk, associated with metformin use in subjects with T2D and COVID-19, might be generalizable. Furthermore, these findings underline the importance of following general diabetes treatment and prevention guidelines and not delaying any metformin treatment. Especially during this pandemic that puts subjects with diabetes at particularly high risk, this treatment might not only help with diabetes management, but also reduce the risk of adverse outcome in case of a COVID-19 infection.

At this point, the mechanisms by which metformin might improve prognosis in the context of COVID-19 are not known. Our findings suggest that they go beyond any expected improvement in glycemic control or obesity as HbA1C or BMI were not lower in COVID-19 survivors on metformin. Interestingly, metformin has previously been shown to also have anti-inflammatory (15, 16) and anti-thrombotic effects (17, 18) and excessive inflammatory responses, e.g., cytokine storm as well as disseminated thromboembolic events have been recognized as deadly complications of COVID-19 infection (19–21). It is therefore tempting to speculate that by exerting some of its anti-fibrinolytic activities (17) and inhibiting inflammatory cytokines such as tumor necrosis factor alpha or interleukin-6 (15, 16), suspected to play a role in the immune response to COVID-19 (14), metformin might improve outcome. In fact, even prior to the COVID-19 pandemic, preadmission metformin use was found to be associated with reduced mortality in medical and surgical intensive care patients with T2D (22).

While diabetes has been recognized universally as one of the major comorbidities adversely affecting COVID-19 outcome, the factors responsible for this phenomenon are not well understood. Of note, we found that the increased mortality risk of subjects with diabetes persisted even after correcting for covariates such as age, race, obesity and hypertension, suggesting that while these factors might contribute to a worse outcome, they cannot fully account for it. Also, long-term glycemic control as assessed by HbA1C did not affect mortality in our study, in alignment with previous reports (6). Similar to the issue with metformin, other factors such as diabetes-associated inflammation (23) and coagulopathy (24) may therefore play a more prominent role in this regard. In addition, a recent report also demonstrated that pancreatic beta cells can get infected and damaged by SARS-CoV-2 (25) providing a potential explanation for the extremely high insulin requirements seen in some subject with COVID-19 as well as the development of diabetic ketoacidosis and possibly new onset diabetes (26, 27).

Higher age and male sex were the other independent risk factors associated with increased mortality that we found consistently across subjects with and without diabetes. In fact, the mortality rate in males was more than 2-fold higher than in females, which is in line with previous studies (28). Many theories have been proposed for why this might be, including the different concentrations of sex steroids, different fat distribution, different level of circulating pro-inflammatory cytokines and different innate and adaptive immune response to viral infections (28, 29). In fact, due to this striking sexual dimorphism, studies using anti-androgens in COVID- 19 positive men are currently ongoing.

In our cohort being African-American appeared to be primarily a risk factor for contracting COVID-19 rather than for mortality. These findings are supported by a recent study using an integrated-delivery health system cohort with similar demographics (∼30% Blacks/African-American), which found that Black race was not associated with higher in-hospital mortality than White race. This suggests that any racial disparity observed may be more likely due to exposure risk and external, socioeconomic factors than to biological differences. The fact that other geographic areas (mostly with a smaller proportion of African-Americans), did see a difference in mortality (10), might be related to issues with healthcare access.

Taken together, our study reaffirmed the role of the major comorbidities associated with COVID-19 in a more diverse population with a higher proportion of African-Americans, demonstrated the prominence of diabetes as an independent risk factor associated with higher mortality and revealed that metformin use prior to a diagnosis of COVID-19 was associated with a consistent and robust decrease in mortality in subjects with diabetes. Future studies will have to explore how metformin might confer these protective effects, provide a careful risk benefit assessment and determine whether the indications for metformin treatment should be broadened in the face of the ongoing COVID-19 pandemic.

## Data Availability

N/A

## AUTHOR CONTRIBUTIONS

A.B.C and T.G. were responsible for data acquisition and analysis, P.L. performed all the statistical analyses, M.M. and F.O. helped with the approach and interpretation, A.S. conceived the study and wrote the manuscript. All authors have read and approved the paper.

## ACKNOWLEDGMENTS

A.S. is supported by National Institutes of Health grants R01DK078752 and U01DK120379 and the UAB Center for Clinical and Translational Science (CCTS) by UL1TR001417.

